# Differential Deep Brain Stimulation Sites and Networks for Cervical vs. Generalized Dystonia

**DOI:** 10.1101/2021.07.28.21261289

**Authors:** Andreas Horn, Martin Reich, Siobhan Ewert, Ningfei Li, Bassam Al-Fatly, Florian Lange, Jonas Roothans, Simon Oxenford, Isabel Horn, Steffen Paschen, Joachim Runge, Fritz Wodarg, Karsten Witt, Robert C. Nickl, Matthias Wittstock, Gerd-Helge Schneider, Philipp Mahlknecht, Werner Poewe, Wilhelm Eisner, Ann-Kristin Helmers, Cordula Matthies, Joachim K. Krauss, Günther Deuschl, Jens Volkmann, Andrea Kühn

## Abstract

Dystonia is a debilitating disease with few conservative treatment options but many types of isolated dystonia can be effectively treated using deep brain stimulation (DBS) to the internal pallidum.

While cervical and generalized forms of isolated dystonia have been targeted with a common approach to the posterior third of the nucleus, large-scale investigations between optimal stimulation sites and potential network effects in the two types of dystonia have not been carried out.

Here, we retrospectively investigate clinical results following DBS for cervical and generalized dystonia in a multi-center cohort of 80 patients. We model DBS electrode placement based on pre- and postoperative imaging and introduce a novel approach to map optimal stimulation sites to anatomical space. Second, we analyse stimulation in context of a detailed pathway model of the subcortex to investigate the modulation of which tracts accounts for optimal clinical improvements. Third, we investigate stimulation in context of a broad-lense whole-brain functional connectome to illustrate potential multisynaptic network effects. Finally, we construct a joint model using local, tract- and network-based effects to explain variance in clinical outcomes in cervical and generalized dystonia.

Our results show marked differences in optimal stimulation sites that map to the somatotopic structure of the internal pallidum. We further highlight that modulation of the pallidofugal main axis of the basal ganglia may be optimal for treatment of cervical dystonia, while pallidothalamic bundles account for effects in generalized dystonia. Finally, we show a common multisynaptic network substrate for both phenotypes in form of connectivity to cerebellum and somatomotor cortex.

Our results suggest a multi-level model that could account for effectivity of treatment in cervical and generalized dystonia and could potentially help guide DBS programming and surgery, in the future.

## Introduction

Deep brain stimulation in patients with treatment-refractory idiopathic dystonia is a well-established therapy with excellent short- and long-term clinical results ^1–4^. However, in the only controlled trial ^1^ and its open 5-year follow-up ^2^, as well as uncontrolled trials with blinded observers ^5^, around 25% of patients had poor response, which was a primary stimulus for the present work. Moreover, while targeting the internal pallidum (GPi) has been successful, there is still a gap in our understanding of which specific sites within the nucleus lead to network modulation of i) localized tracts and ii) whole-brain functional networks. Finally, whether targeting could be refined for cervical vs. generalized dystonia has not been investigated in large cohorts, so far.

Here, we re-revisit a particularly large multi-center cohort ^6^ with the aim to relate treatment effects to connectional concepts and to investigate potential differences in treatment response of cervical vs. generalized dystonia patients. We do so by introducing a novel sweetspot mapping method that is based on electric fields rather than binarized volumes of tissue activated, as well as the recently introduced DBS fiber filtering ^7,8^ and DBS network mapping ^9,10^ approaches.

Hypotheses for this study were established based on two lines of reasoning. The first involves somatotopic organization of the GPi with neurons responding to the orofacial, forelimb, and hindlimb regions of primary motor cortex located along the ventral-to-dorsal axis in its posterolateral part ^11–13^. Hence, potentially, ventral stimulation sites could be more specific for responders in cervical dystonia with generalized dystonia optimally responding to a larger or more diffuse stimulation territory. Second, we developed one hypothesis based on the microanatomy of the GPi, which involves that two streams of fibers pass the GPi in largely orthogonal fashion to one another ^14,15^. First, there is the extension of the striatopallidofugal system in form of Edinger’s comb (connecting striatum and pallidum to SNr and STN). Second, there are the pallidothalamic projections (in form of ansa and fasciculus lenticulares). We aimed to investigate differential effects by leveraging group cohort data of stimulation sites. A more detailed anatomical discussion that led to this hypothesis is given in the supplementary material and summarized in **figure 1**.

**Figure 1:**
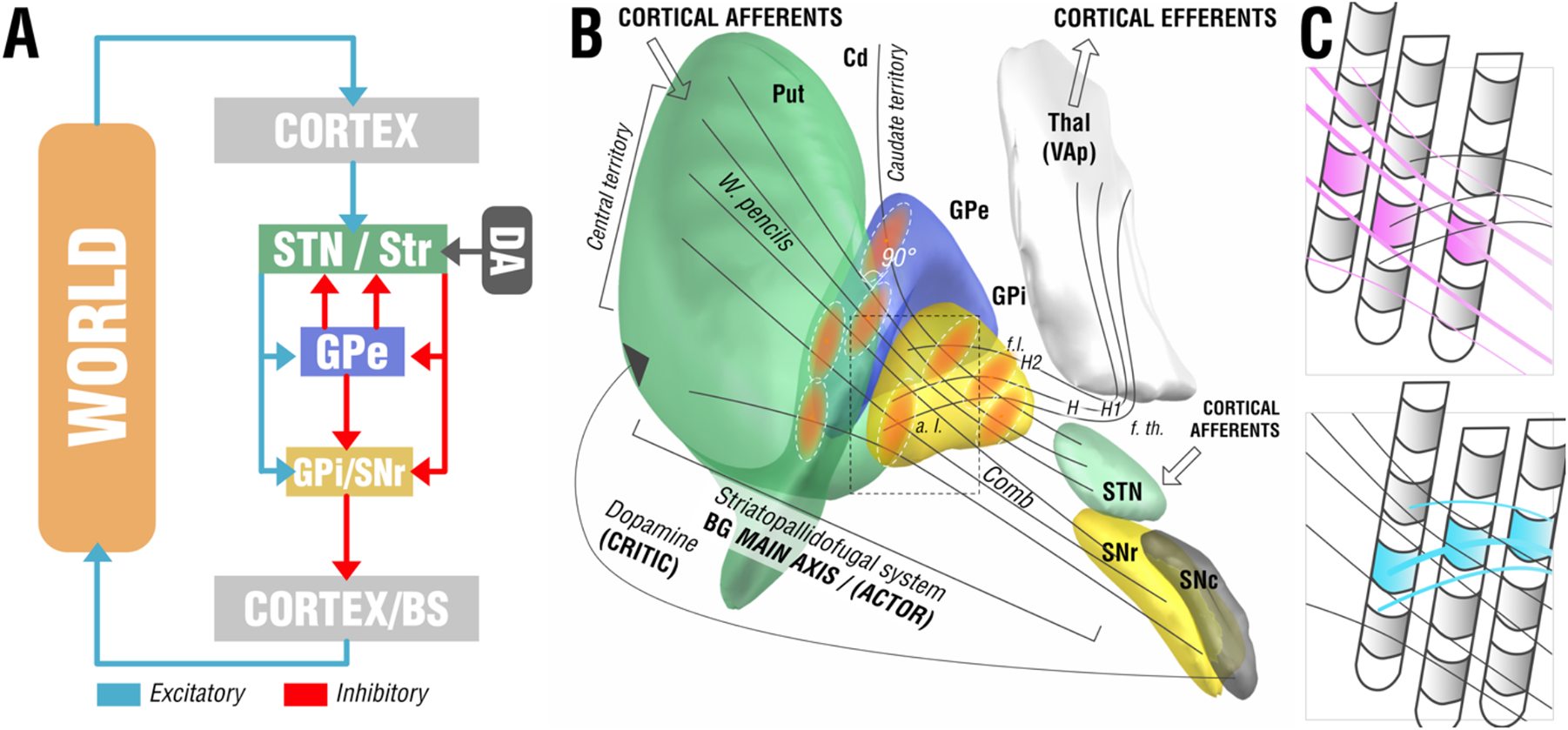
Functional-anatomical model leading to the core hypothesis for the present study. A) Basal-ganglia model in context of a reinforcement-learning context adapted from ^16^. The left panel shows the main axis of the basal ganglia (actor) with a three-layer model in which both striatum and subthalamic nucleus form entry nodes and GPi (I) and substantia nigra pars reticularis (R) serve as output ganglia, feeding information back (via the thalamus) to the cortex and passing it on to brainstem centers (BS). Dopaminergic input serves as one of multiple critics to reinforce successful motor behavior. B) Translation of the model to the anatomical domain based on information shown in figure S1. The striatopallidofugal system and pallidothalamic fibers serve as the main axis (actor) and receive feedback from dopaminergic centers, especially substantia nigra pars compacta (SNc). Pallidal receptive fields reside in a 90° angle to the striatopallidofugal fiber system, pallidothalamic output tracts traverse the main axis in equally orthogonal fashion. C) Hypothesis generation for the present study based on anatomical considerations. Two scenarios are possible (shown as cut-out box from panel B). Top: Active contacts (pink) of top-responding patients are located along the direction of striatopallidonigral fibers. In this case, our results would reveal activation of these fibers to best account for clinical outcome. Bottom: Instead, active contacts (cyan) could also be located along pallidothalamic tracts (ansa lenticularis; a.l. and fasciculus lenticularis; f.l.). In this case, our results would reveal activation of these fibers to best account for clinical outcomes.

From this concept, we derived two competing hypotheses that are illustrated in **Figure 1** C. We registered all DBS electrodes and stimulation volumes to a common space model of the basal ganglia in which the anatomical fiber projections were informed by a recently published and highly accurate pathway atlas of the basal ganglia ^17^. We hypothesized that three testable scenarios could be present in the data. First, the stimulation volumes of top-responding patients in our sample could be arranged in a way that would not allow any anatomical conclusions (i.e., in a random fashion throughout the cohort). This would favor a null hypothesis according to which our data would not be able to differentiate between the two fiber systems. Alternatively, stimulation volumes from top-responding patients could be arranged in a *radial* way – along the fibers of the *striatopallidofugal system* – or in an *orthogonal* way – along the *pallidothalamic system*. In case one of those scenarios would hold true, our data could associate one of the two fiber systems with optimal clinical outcomes.

Here, we aimed at addressing this question using the DBS fiber filtering method to isolate tracts that are predominantly associated with top-responding patients in cervical vs. generalized dystonia. The method was introduced in rudimentary form in 2019 ^7^ and has been subsequently refined ^8,18^. We complemented the approach by a novel sweet-spot mapping algorithm that directly works on electric fields instead of binarized stimulation volumes. Here, the aim was to map optimal stimulation sites to somatotopic regions within the GPi (see *methods: Modeling considerations*). Finally, to complement results with a “broad-lense view” that would include polysynaptic networks, we applied the DBS network mapping approach to identify whole-brain functional networks that accounted for optimal treatment response ^9^.

## Methods

### Patient Cohorts and Imaging

Eighty DBS patients from five different centers were retrospectively included in this study after meticulous inspection of imaging quality (25 patients were excluded due to poor imaging quality after visual inspection). All patients underwent DBS surgery for either cervical (N = 46) or generalized (N = 34) dystonia and received 2 quadripolar DBS electrodes (either model 3389 or 3387; Medtronic, Minneapolis, MN). The surgical procedure was similar in all centres, and has been described previously ^19^. The neurostimulation parameters were programmed according to best clinical practice by the local DBS neurologist, based on clinical response testing. All video sequences were rated retrospectively by the same movement disorder neurologist (M.R.), using either the Toronto Western Spasmodic Torticollis Rating Scale (TWSTRS) in subjects with cervical dystonia or the Burke-Fahn-Marsden Dystonia Rating Scale (BFMDRS) in patients with generalized or segmental dystonia (in the following referred to as generalized dysonia). Results were normalized by calculating the percentage change of the TWSTRS and the BFMDRS. In subjects with cervical dystonia, the TWSTRS motor score improvement without using the duration factor (item Ib) was assigned to both hemispheres equally; this modified motor score was chosen because the total TWSTRS motor score is too strongly weighted by the duration factor with respect to the improvement of dystonic postures ^19^. In subjects with generalized or segmental dystonia, the global improvement in BFMDRS was associated with the stimulation of both hemispheres. All patients received preoperative MRI and neuropsychological testing to exclude structural or severe psychiatric comorbidities. After surgery, patients received postoperative MRI or CT imaging to confirm electrode placement. The study was carried out in accordance with the Declaration of Helsinki and approved by the institutional review board of the University Hospital of Würzburg (registration no. 150/15).

### DBS electrode localizations and E-field modeling

DBS electrodes were localized using the advanced processing pipeline ^20^ in Lead-DBS (<u>lead-dbs.org</u>; ^21^; RRID:SCR_002915). In short, postoperative CT or MRI were linearly coregistered to preoperative MRI using advanced normalization tools (ANTs; <u>stnava.github.io/ANTs/</u>; ^22^). Coregistrations were inspected and refined if needed. A brain shift correction step was applied as implemented in Lead-DBS. All preoperative volumes were used to estimate a precise multispectral normalization to ICBM 2009b NLIN asymmetric (“MNI”) space ^23^ applying the ANTs SyN Diffeomorphic Mapping ^24^ using the preset “effective: low variance default + subcortical refinement” in Lead-DBS. This approach was top-performer to segment the GPi with precision comparable to manual expert segmentations in a recent comparative study ^25^ which was further replicated by a different group ^26^. Normalization warp-fields were further manually adapted using the respective module in Lead-DBS ^27^, as well as an unpublished reiteration of the approach, termed “WarpDrive” that will be published elsewhere. DBS contacts were automatically pre-reconstructed using the phantom-validated and fully-automated PaCER method ^28^ or the TRAC/CORE approach ^21^ and manually refined if needed. Atlas segmentations in this manuscript are defined by the DISTAL atlas ^29^. Group visualizations were performed using the Lead group toolbox ^30^.

Electric fields (E-fields) were estimated in native space based on the long-term DBS settings applied using an adaptation of the SimBio/FieldTrip pipeline ^31^ as implemented in Lead-DBS ^20^. Briefly, using the finite element method, the static formulation of the Laplace equation was solved on a discretized domain represented by a tetrahedral four-compartment mesh (composed of gray and white matter, metal, and insulating electrode parts). Electric fields were transformed to MNI space using the same refined normalization warpfields described above. Since no lateralized effects were expected ^6^, for all subsequent analyses, E-fields were nonlinearly flipped to the other hemisphere in order to overlay 2 × 80 = 160 E-fields across the whole cohort.

### Modeling considerations

Estimated after ^32^, each cubic millimeter of cortex is filled with ∼170,000 neurons, each with an average number of ∼10,000 inputs and outputs. According to numbers aggregated by Bergman ^33^, the GPi is less densely populated, with only ∼1,000 neurons per cubic millimeter. For axonal numbers, following ^34^, each fiber bundle in a standard neuroimaging analysis represents 10^3^-10^5^ tightly packed axons. Many DBS studies aimed at modeling discretized and realistic axonal cable models, in the past ^35–37^. However, given these sheer numbers of axons involved, here, we chose to assume probabilistic axonal *populations* in each brain voxel and represented by each fiber tract, instead of modeling representative *single axons*. Such populations will have more diffuse firing properties that could encode numeric variables, rather than following an all-or-nothing firing property that would be assumed for single axons ^38^. While single axons fire in an all-or-nothing fashion, activations of larger axonal populations within a voxel may be better represented by a probabilistic fashion which is dependent on the applied voltage ^39–41^. In other words, on a population level, the “degree” of activation will be stronger under higher voltages applied or when closer to the electrodes. Crucially, there is large amount of uncertainty about the exact *relationship* between voltage and axonal firing that needs patient-specific calibration even when applying more realistic biophysical models (Howell *et al*., 2021). Since this relationship is unclear, we applied Spearman’s rank correlations in our sweetspot and fiber filtering models. We believe that this simple model could have a crucial advantage, since it would show maximal values (R → 1.0) for any type of function that is monotonically increasing. In other words, the concept could be robust toward the exact relationship (be it linear, cubic or logistic) between amplitude and modulation.

### DBS Sweetspot Mapping

#### Model (Figure 2 A)

Using E-fields calculated in each patient, and the aforementioned considerations in mind, a novel approach to define optimal stimulation sites was applied (**Figure** 2 A), which was inspired by the DBS network mapping approach introduced earlier (also see below; ^9^). E-fields represent the first derivative of the estimated voltage applied to voxels in space and their vector magnitudes are hence stronger in proximity of active electrode contacts with a rapid decay over distance. For each voxel covered by the group of E-fields across the cohort in MNI space, E-field vector magnitudes across patients were Spearman rank correlated with clinical improvements. Since not all voxels were covered by the same amount of E-Fields, the area of interest was restricted to voxels that were at least covered by 30% of E-Fields with a vector magnitude above 150 V/m, which is around a typical value that has been assumed to activate axons ^42^. The resulting sweetspot maps would peak at voxels in which stronger E-fields were associated with better treatment responses. The map would have negative values for voxels with the opposite relationship.

**Figure 2:**
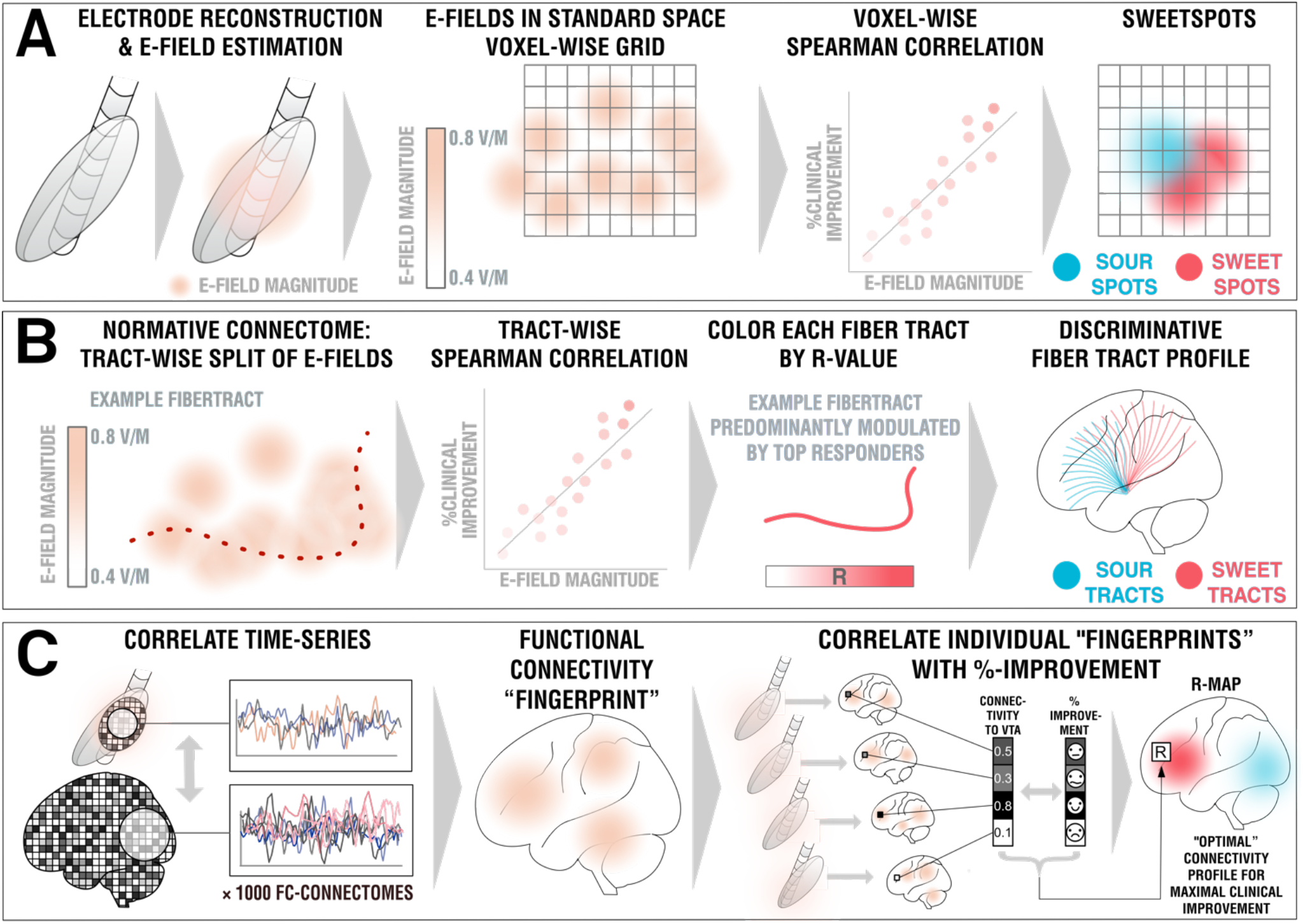
Overview about the three methods applied. A) DBS Sweetspot mapping. Based on DBS electrode localizations carried out with Lead-DBS, electric fields (E-Fields) were estimated using a finite element approach based on the long-term stimulation parameters applied in each patient. E-Fields were then warped into MNI space. For each voxel, the E-Field vector magnitudes and clinical improvements were rank-correlated, leading to a map with positive and negative associations (sweet and sour spots). B) DBS fiber filtering. Again, E-Fields were pooled in standard space and the group was set into relationship with all of 26,800 tracts forming a predefined set of normative pathways ^17^. Sum E-Field magnitudes along each tract were aggregated for each patient and again rank-correlated with clinical improvements, attributing positive vs. negative weights to each tract (sweet and sour tracts). C) DBS network mapping. Seeding BOLD-signals from each E-Field in a database of 1,000 healthy brains led to functional connectivity maps that were averaged to form a functional connectivity “fingerprint” for each patient. Voxels in these were correlated with clinical improvements to create an R-map model of optimal network connectivity.

#### Estimates

Mutliplying each voxel of a single E-field with the resulting sweetspot map and calculating the sum across voxels led to estimates of how a specific E-field would perform (i.e., estimates of clinical improvements following DBS). If the E-field peaked at similar locations as the sweetspot map, a high estimate would result. If it would peak at a valley of the map, low or even negative estimates would result.

### DBS Fiber Filtering

#### Model (Figure 2 B)

For a finite set of 28,600 subcortical fibertracts represented within the Basal Ganglia Pathway Atlas ^17^ and each E-field in each patient, a value of probabilistic impact on the tract was calculated by summing the E-field magnitude vectors along the tract. This led to a matrix of 28,600 × 160 dimension, each entry denoting the sum “impact” of each E-field on each tract. Again, the exact relationship between E-field magnitude and activations of axonal populations is dependent on multiple of factors unknown in the individual patient (axonal shape, diameters, myelinisation, degrees of arborization of both dendritic and axonal terminals, numbers of nodes of Ranvier, conductivity of axonal, interstitial vs. myelin components, degree of microstructural anisotropy, heterogeneity and dispersivity of tissue conductivity, specific properties of the encapsulation layer, capacitive properties, etc.). Hence, again, Spearman’s rank correlations were chosen which would account for any type of monotonically increasing function. This led to a model of 28,600 correlation coefficients (one for each tract), showing positive values for tract populations maximally “impacted” by electrodes in top responding patients and negative values for the ones preferentially modulated in poor responding patients.

#### Estimates

In a similar fashion, single E-fields were probed based on the estimated tract model. If their “peaks” resided on positively weighted tracts and their “valleys” on negatively (or less positively) weighted tracts, they received a high score estimate. Again, the exact (linear or nonlinear) relationship remains elusive; so, a third time, Spearman’s rank correlations were applied, again.

### DBS Network Mapping

#### Model (Figure 2 C)

In a last approach, we calculated whole-brain functional connectivity estimates seeding from E-fields based on a library of resting-state functional MRI (rs-fMRI) scans acquired in 1,000 healthy participants ^43,44^, following the approach of ^9^. This method allowed to investigate the functional connectivity profile of a specific DBS electrode *within the average human brain* – and the resulting maps have been termed *connectivity fingerprints*, in the past ^10^. In analogy to the sweetspot model, voxel-wise correlations between Fisher-z-scored connectivity strengths and clinical improvements were calculated, which led to R-map models of optimal connectivity. Here, Pearson’s correlations were applied since underlying values are normal distributed and linear relationships could be assumed (in comparison, E-fields used above are composed of highly skewed distributions). As for sweetspot and tract filtering models, one DBS network mapping model was calculated for cervical and generalized dystonia cases, separately. However, given the more “broad-lense” view these models impose, one additional model was calculated on the entire cohort. Finally, following the approach of ^10^, an agreement map was calculated between cervical and generalized models, which retained only voxels that had the same sign in both models (and multiplicated their absolute values). The latter was performed to identify potential *common denominators* in network effects across cervical and general dystonia types.

#### Estimates

Spatial similarities between single connectivity fingerprints and R-map models were calculated using voxel-wise spatial correlations. This led to positive high correlation values for cases in which fingerprints graphically matched the (optimal) connectivity profile represented by the R-map model – and lower or even negative values for other cases.

## Results

Electrodes were accurately placed within the target region for all patients with active contacts within or close to the GPi (**Figure** 3). Clinical results of this retrospective cohort are described in more detail, elsewhere ^6^. Briefly, our DBS cohort included 80 patients operated at five different DBS centers (34 female, mean age 48.3 ± 16.0 years), 43 with cervical and 37 with generalized dystonia (see Table 1).

**Figure 3:**
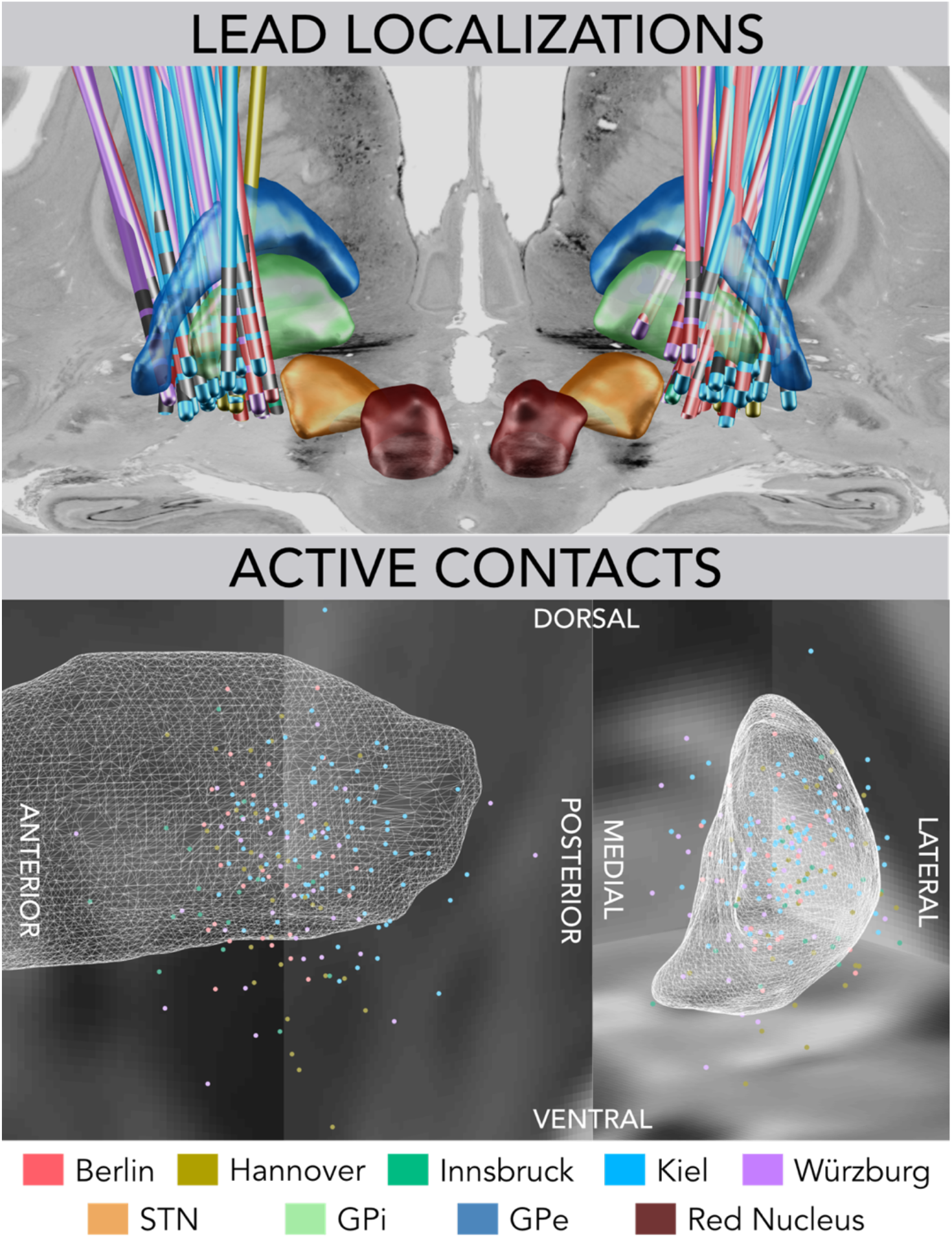
Reconstructions of DBS electrode placement of the all five cohorts color-coded by center (top). Active DBS contacts of the group localized well to the posteriolateral portion of the GPi (white wireframes) which corresponds to its sensorimotor functional zone.

**Figure 4:**
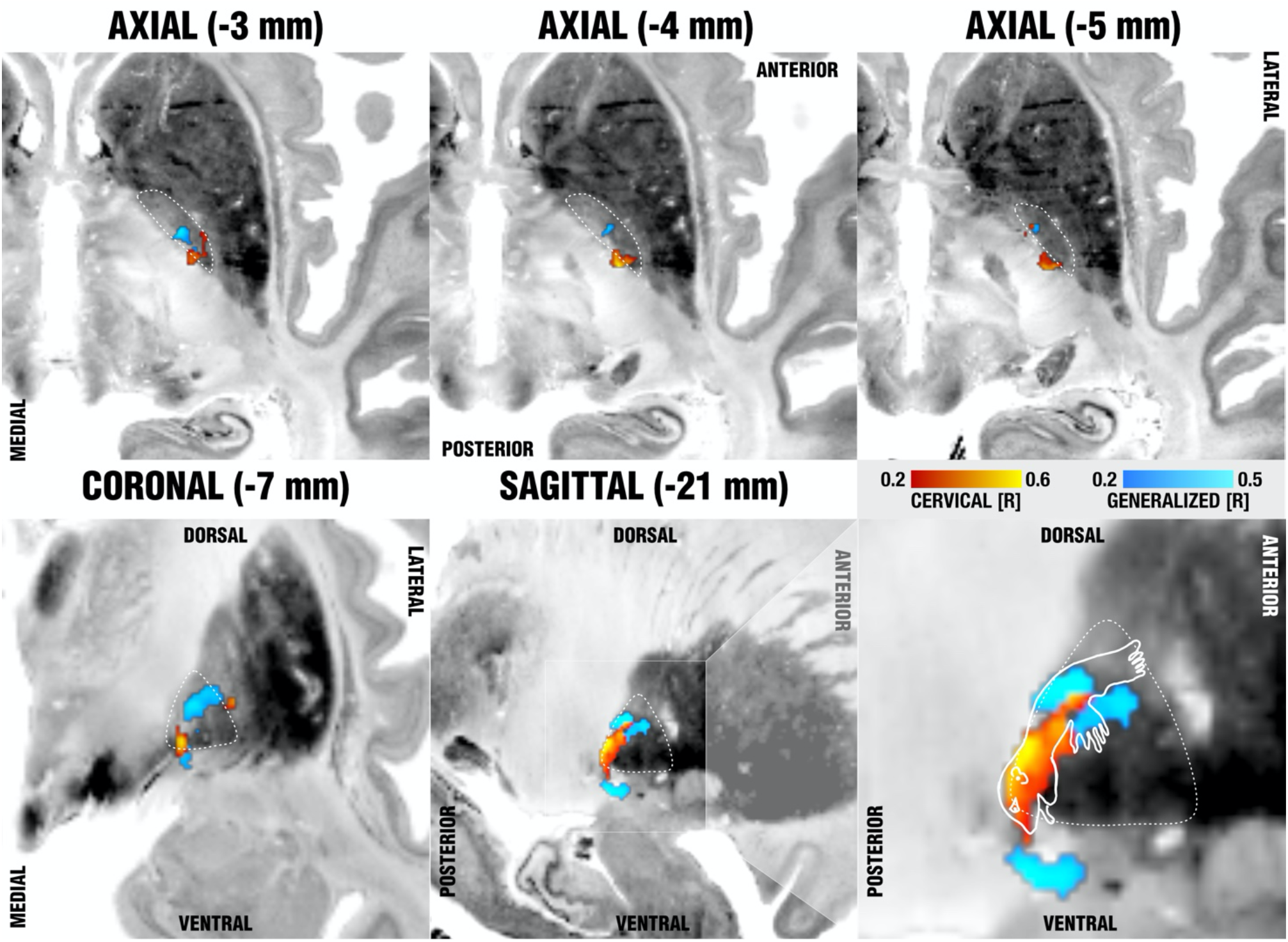
Sweetspot mapping of cervical (red) vs. generalized (blue) subcohorts matches somatotopic organization of the GPi as defined by ^12^. Voxels are color-coded by the degree of correlation between % improvements of either TWSTRS (cervical, hot colors) or BFMDRS (generalized, cool colors) and shown on multiple axial (top) and coronal/sagittal slides (bottom) on top of the BigBrain template ^47^. The last panel shows the homuncular representation of the pallidum following reports by Nambu et al. which stated that neurons responding to the orofacial, forelimb, and hindlimb regions of motor cortex are located along the ventral-to-dorsal axis in the GPi.

**Table 1:** Patient demographics.

| DBS center | Mean Age | N cervical (female) | N generalized (female) | N total (female) | %-Clinical Improvements (cervical) | %-Clinical Improvements (generalized) | %-Clinical Improvements (combined) |
| --- | --- | --- | --- | --- | --- | --- | --- |
| Berlin | $51.4 \pm 16.7$ | 4 (2) | 6 (1) | 10 (3) | $28.6 \pm 50.8$ | $51.3 \pm 28.8$ | $42.2 \pm 38.2$ |
| Hannover | $47.9 \pm 17.8$ | 6 (2) | 3 (2) | 9 (4) | $48.7 \pm 42.6$ | $36.1 \pm 44.1$ | $44.5 \pm 40.7$ |
| Innsbruck | $46.3 \pm 15.2$ | 5 (3) | 2 (0) | 7 (3) | $49.4 \pm 22.3$ | $73.9 \pm 22.9$ | $58.8 \pm 24.8$ |
| Kiel | $47.0 \pm 15.3$ | 21 (13) | 20 (7) | 41 (20) | $66.0 \pm 27.1$ | $73.9 \pm 22.9$ | $69.9 \pm 25.2$ |
| Würzburg | $50.8 \pm 18.6$ | 7 (4) | 6 (4) | 13 (4) | $48.0 \pm 31.9$ | $87.7 \pm 11.3$ | $66.3 \pm 31.4$ |
| **Total** | $48.3 \pm 16.0$ | 43 (24) | 37 (14) | 80 (34) | $55.2 \pm 33.0$ | $69.9 \pm 27.3$ | $62.0 \pm 31.2$ |

On a local level (DBS sweetspot mapping), voxels in the posterior ventromedial GPi were associated with optimal improvements of the cervical cohort, whereas voxels equally medial but at a slightly more anterior and dorsal subregion of the GPi were most associated with improvements in the cohort with generalized dystonia. The cervical sweetspot map peaked at ±20.4, -12.4, z = -5.2 mm (MNI coordinates; with a Spearman’s rank correlation coefficient of R = 0.58), which was located precisely at the medial pallidal border and a mere 2.6 mm apart from the maximal sweetspot coordinate reported by Reich et al. (±19.4,-10.1,-5.9 mm) ^6^. This is important given their spot was calculated with a completely different methodological pipeline. Similarly, the spot precisely matched the finding by Mahlknecht and colleagues in cervical dystonia ^45^. The generalized sweetspot map peaked at ±21.1, -9.1, z = -0.14 mm (R = 0.67), i.e., more dorsal, and anterior and about 6 mm apart from both the cervical spot and the optimal coordinate reported by Reich et al. When visualized in context of the GPi, cervical sweet spot regions localized to the cervical somatotopic motor region of the pallidum as described by ^12^, which map to the ventral border of the pallidum. Generalized sweet spot regions were more outstreched, potentially incorporating a larger somatotopic fraction of the motor pallidum (see last panel of Figure 5, which summarizes sweetspot results). The exact peaks of this spot resided in two sites, dorso-anterior and ventral to the cervical peak, which could potentially associate with course of the ansa lenticularis, which has been described to course ventrally to the pallidum ^6,46^. Beyond this ventral site, in synopsis with a homuncular projection adapted from ^12^, the largest peak resided within the hand and trunk region of the pallidum (figure 5).

**Figure 5:**
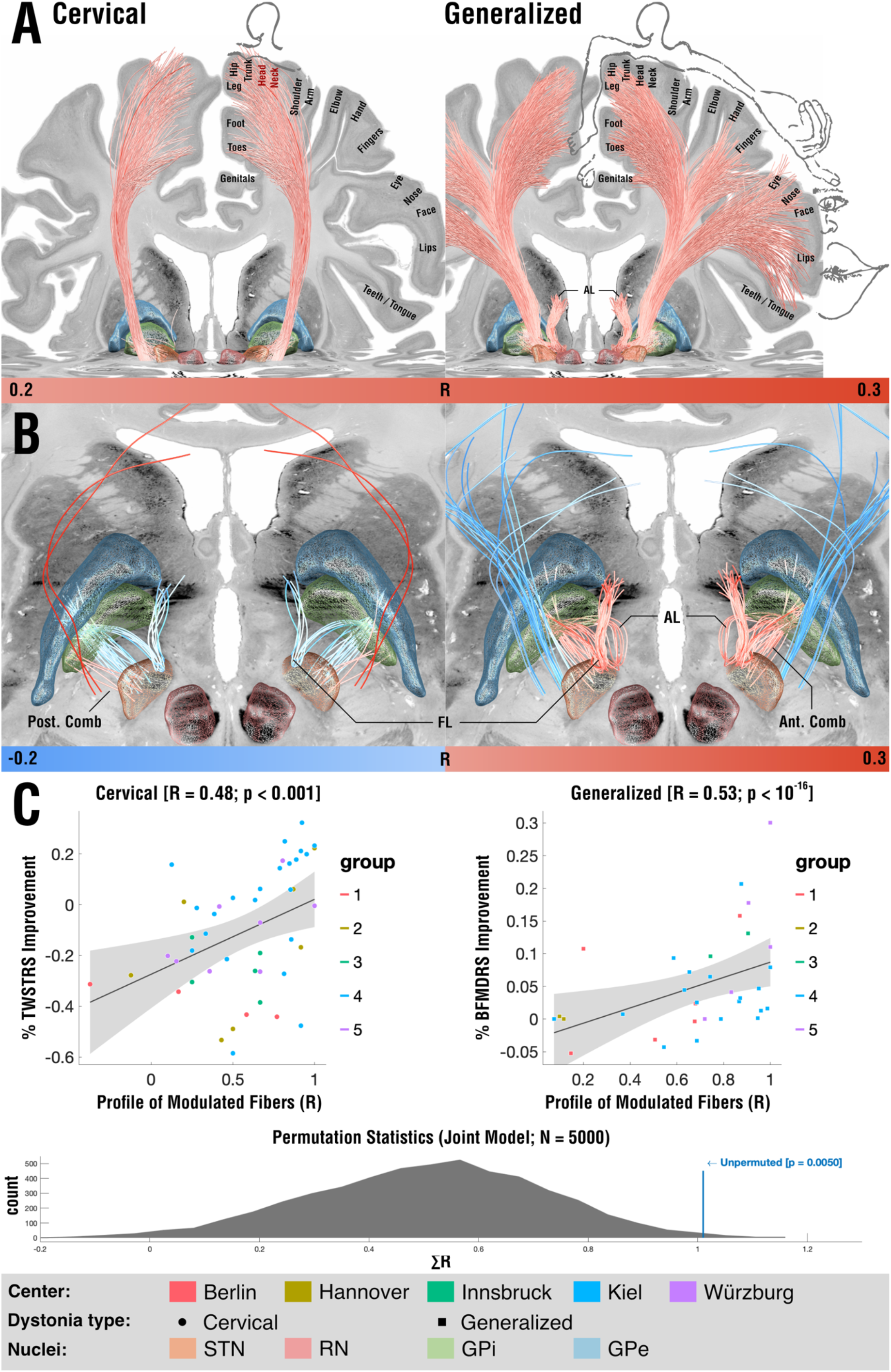
Tracts associated with optimal outcome for patients with cervical (left) and generalized (right) dystonia. A) On a broader scale (slightly lower threshold), modulation of corticofugal tracts from the somatomotor head & neck region was associated with optimal outcomes in cervical dystonia, while tracts from the whole somatotopical domain with generalized dystonia. B) On a localized level (slightly higher threshold), in cervical dystonia, pallidofugal tracts of the posterior comb system were associated with optimal outcomes. In contrast, fibers from the fasciculus lenticularis were negatively associated. In generalized dystonia, both pallidothalamic bundles (ansa and fasciculus lenticularis) were associated with optimal outcomes, as was a more anterior portion of the comb system.

On a tract level, from all 28,600 fiber bundles included within the Basal Ganglia Pathway atlas ^17^, the clinical outcomes in the *cervical* cohort correlated most with i) pallidosubthalamic fibers in the posterior (i.e., motor) part of Edinger’s comb system and ii) indeed, corticospinal fibers of passage connecting to the head and neck region of the sensorimotor cortex. Indeed, this finding is in agreement with the medial position of sweetspots identified in the present study and the one by Reich and colleagues ^6^. We must emphasize that methodological constraints hinder us from concluding with certainty, whether in the actual brain of patients, these tracts would indeed map to i) fibers of passage, ii) corticopallidal tracts (which are sparse but present), iii) peri-pallidal projections to cortex as described by Parent et al. ^48^ or iv) corticospinal/-pontine projections. Crucially, in the cervical cohort, fasciculus lenticularis and a more anterior part of Edinger’s comb (still within its motor domain) were negatively associated with optimal clinical outcomes. In the cohort with *generalized* dystonia, tracts most associated with optimal outcomes were the pallidothalamic tracts, i.e., fasciculus and ansa lenticulares, as well as some of the more anteriorly situated comb fibers. Instead, some even more medially located fibers of passage within the internal capsule were *negatively* associated with optimal outcomes. Figure 5B summarizes these results. When lowering the visualization threshold to allow for a more broad-lense view on involved networks, tracts associated with optimal outcomes in cervical dystonia involved the cortical connections to the head/neck region of the somatomotor cortex, while regions to the full somatotopic spectrum were associated with positive outcomes in the generalized dystonia cohort (figure 5A). The two models explained ∼23% (R = 0.48; p < 0.001) and ∼28% (R = 0.53; p < 10^−16^) of variance within the whole sample but we must emphasize that this analysis was circular and can merely express the degree of fit between data and model. To account for this, we calculated random permutations (× 5000 iterations) and re-calculated the same model & correlations after permuting improvement values across cohorts. The sum of the two R-values (0.48 + 0.53 = 1.01) was significantly larger in the unpermuted vs. the permuted cases (p = 0.005; figure 5C).

To further extend these insights and to add an additional component to our model, we applied the DBS network mapping approach on estimates of whole-brain functional connectivity as informed by a normative connectome obtained from 1,000 healthy brains. While the structural connectivity estimates from the basal ganglia pathway atlas could investigate, which specific localized connections accounted for clinical outcomes, this additional analysis asked the same question for distributed whole-brain networks that could include indirect, polysynaptic connections, as well. Again, functional connectivity to different sets of regions were associated with optimal outcomes for cervical vs. generalized cohorts, which are summarized in table 2 and shown in figure 6). Most saliently, generalized dystonia was associated with stronger anticorrelations to whole sensory cortices (where in contrast cervical dystonia specifically to the homuncular head/neck regions). Improvements in cervical dystonia were associated with positive connections to SMA and posterior cingulate cortex, while in generalized dystonia, the same was true for ventral ACC and precuneus. When pooling across all patients irrespective of dystonia type (‘Combined’ panel in figure 6), anticorrelations to somatosensory cortex and positive connections to cerebellum, SMA and cingulate cortex were favored. Finally, we calculated an agreement map to visualize regions that positively or negatively correlated in *both* subcohorts alone, which revealed positive connections to cerebellum and anticorrelations to head/neck region of the somatomotor cortex.

**Table 2:**
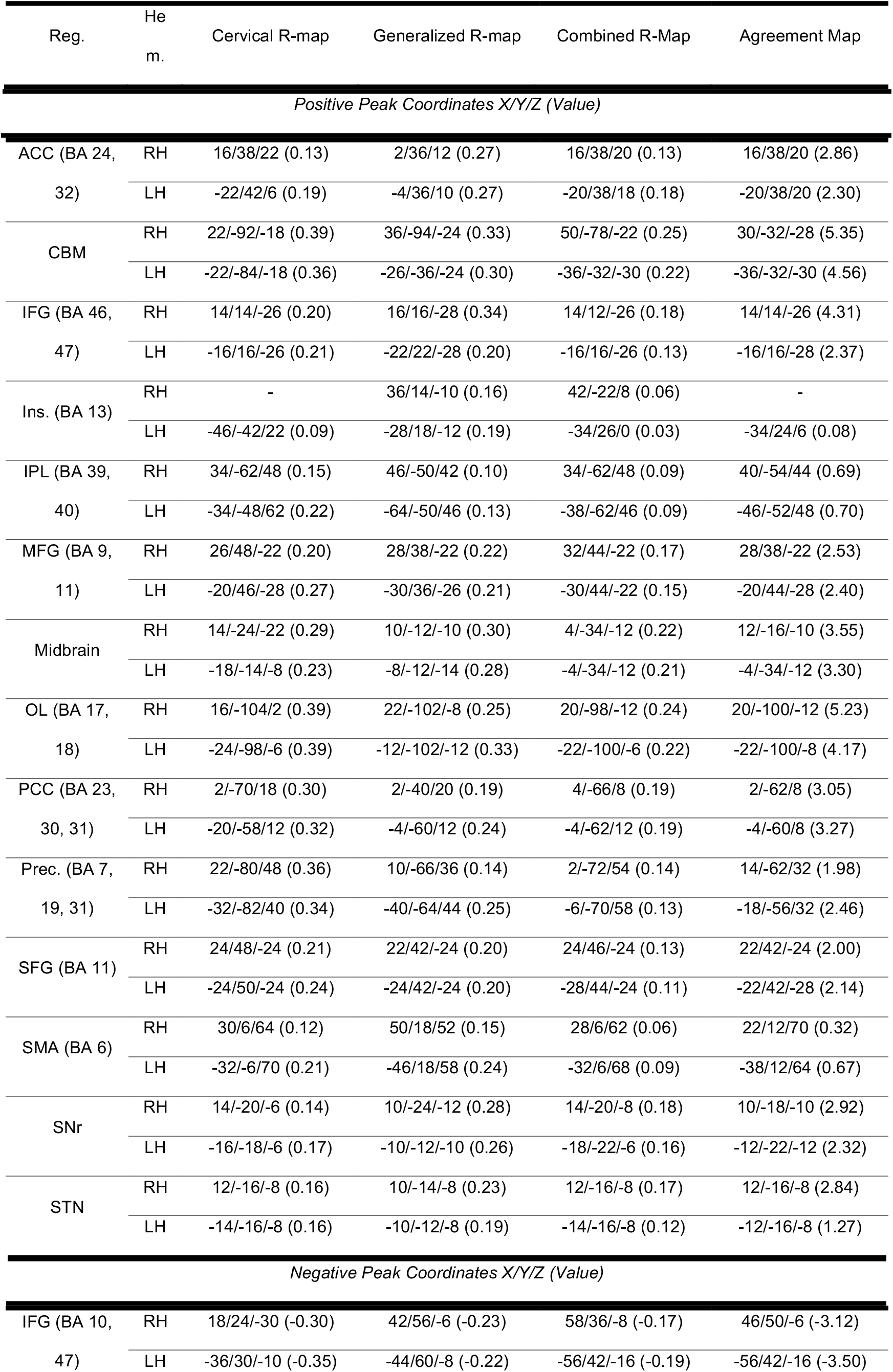
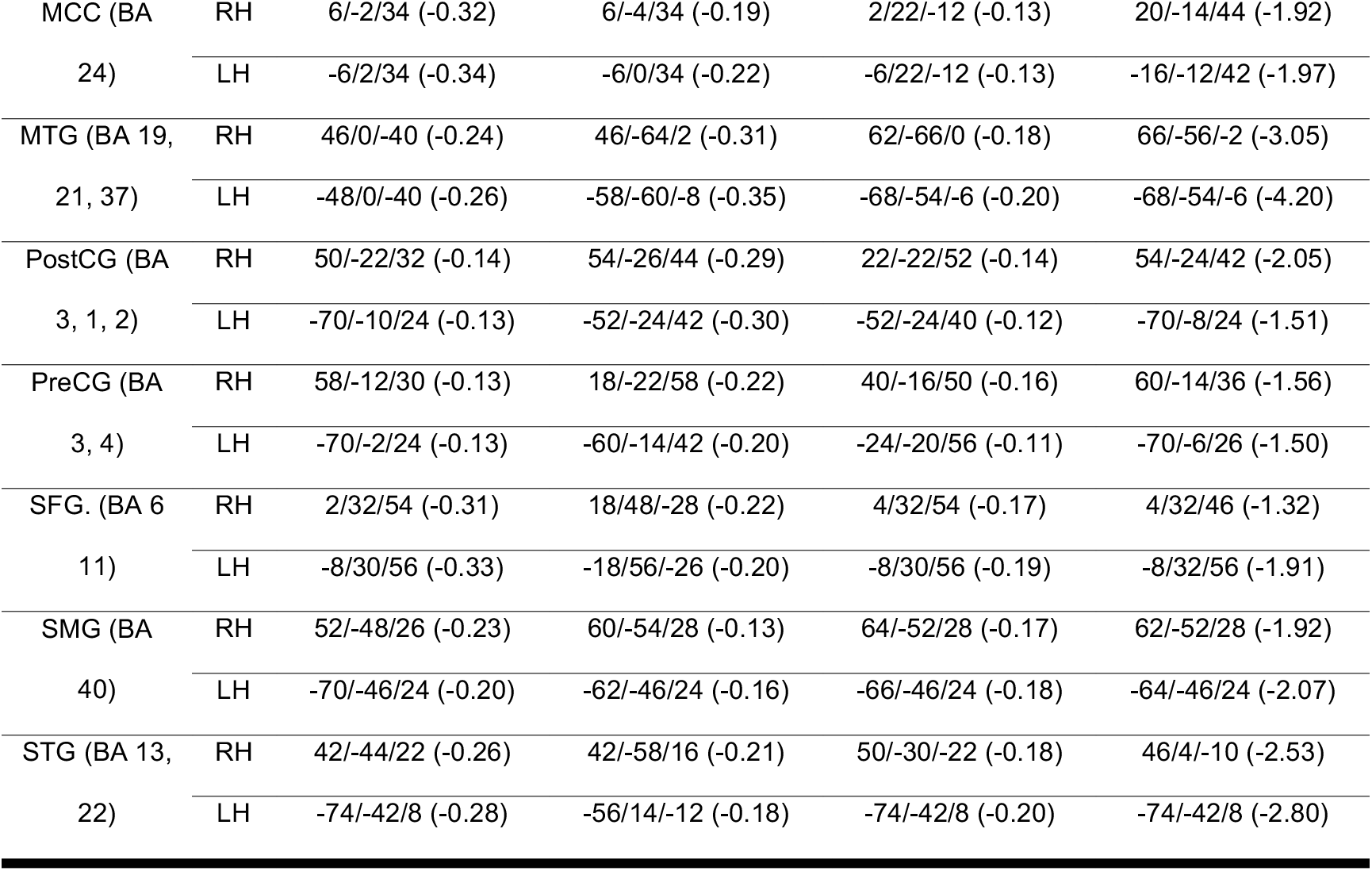
Summary of regions involved in DBS network mapping results. Coordinates of peaks are given in MNI (mm) format with the R-value denoted in brackets. Abbreviations: ACC, anterior cingulate cortex; BA, Brodmann area; CBM, cerebellum; IFG, inferior frontal gyrus, ins., insula; IPL, inferior parietal lobule, MCC, midcingulate cortex; MFG, middle frontal gyrus; MTG, middle temporal gyrus; OL, occipital lobule; PCC, posterior cingulate cortex; PreCG, precentral gyrus; Prec, precuneus; PostCG, postcentral gyrus; SFG, superior frontal gyrus; SMA, supplementary motor area; SMG, supramarginal gurys; SNr, substantia nigra; STG, superior temporal gyrus; STN, subthalamic nucleus

**Figure 6:**
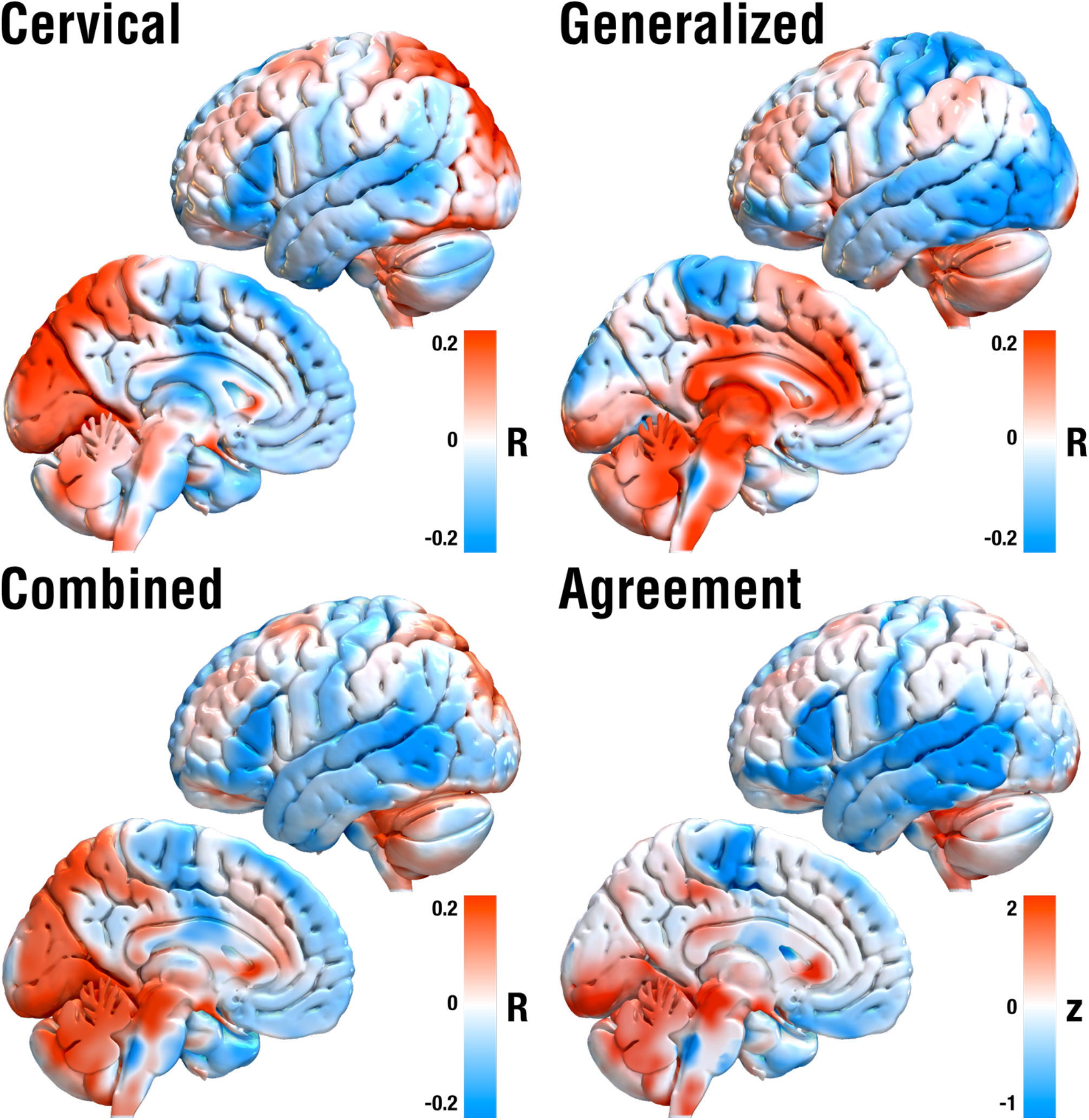
DBS network mapping results based on normative resting-state fMRI data. Red regions show connections positively correlated with clinical improvements, blue regions the opposite. Crucially, optimal networks in cervical vs. generalized dystonia differed substantially but both included positive connections to cerebellum and negative to somatomotor cortices (as revealed by both the combined and agreement maps).

## Discussion

Three main conclusions can be drawn from our study. First, we show first evidence that optimal stimulation sites for cervical and generalized dystonia map to different target regions, tracts, and whole-brain networks. Specifically, our results suggest that optimal stimulation sites within the pallidum map to somatotopic pallidal regions, i.e., the ventral head/neck motor zone of the GPi for maximal benefit in cervical dystonia and a more diffuse mapping to the motor part of the pallidum for generalized dystonia. Second, we show results that suggest specific connections could play a key role in mediating treatment benefit in cervical vs. generalized dystonia. While modulating pallidothalamic tracts accounted for optimal improvements in generalized dystonia, specific corticofugal tracts connecting to head/neck regions of the somatomotor cortex, as well as a specific subpart of pallidosubthalamic connections accounted for effects observed in cervical dystonia. Third, we investigated which whole-brain functional networks would account for optimal treatment success. Analysis again suggested the involvement of differential networks with a common substrate that involved positive connections to cerebellum and negative connections to somatomotor cortex.

As the most salient finding, our report sheds light on a potential segregation between optimal stimulation sites for cervical vs. generalized dystonia at the pallidal level. Namely, stimulation of the pallidofugal bundle was associated with optimal improvement in cervical dystonia, pallidothalamic tracts were with optimal improvements in generalized dystonia. While these systems are segregated, they have a clear common path back to the thalamus and cortex and both coincide with cerebellar input at the thalamic level. We believe these insights could be highly relevant and suitable to form novel hypotheses, but must emphasize potential limitations of the model and techniques (see below) and believe that further confirmation will be mandatory going forward.

### Localized stimulation model

Several optimal stimulation sites within the pallidal region have been suggested for dystonia, in the past. Some have concluded that optimal stimulation sites would be localized in either the intersection between internal and external pallidum ^49,50^. A large study which had analyzed the same sample concluded on a more ventral position ^6^. Here, the focus had been to generate a predictive modeling framework that was able to account for ∼50% of variance in clinical outcomes in out-of-sample data (i.e., patients not seen by the model).

The match between somatotopic pallidal regions and optimal results in cervical and generalized dystonia could be one possible reason for seemingly heterogeneous results in past studies. Regions that did account for optimal outcomes in cervical dystonia indeed precisely mapped to the ventral motor part of the pallidum, as suggested by somatotopic mapping data conducted in primates ^12^. On the other hand, optimal stimulation sites for dystonia comprised vastly spanning parts of the motor pallidum, including the interface between internal and external pallidum (as e.g., reported by ^49^), as well as the portion ventrally to the pallidum reported by ^6^. Hence, somatotopy of the disease could play a major role especially on the dorsoventral axis of the GPi. Needless to say, others have suggested this, before. For instance, Vayassiere et al. concluded that inside the posterolateroventral subvolume of the GPi on the right side, three statistically different locations of electrode contacts were determined to be primary deep brain stimulation treatment sites for particular body parts in cases of dystonia, a notion that our findings confirm ^11^. The novel sweetspot mapping approach we propose here may have the advantage of not being limited to spherical and binary tissue activation models and – at least in theory – would be able to shape out sweet spots of any geometrical form. In combination with the large sample size of our study, the somatotopic results we show could be seen as a useful addition. The regions will be made available under an open license within Lead-DBS software, which could facilitate confirmatory (or directly contradictory) follow-up studies on additional samples.

### Tract-level stimulation model

To expand on the second notion (anatomical considerations), as mentioned in the introduction, the pallidum contains two massive and orthogonal systems of fibers. On one hand, the striatopallidofugal system traverses the pallidum *radially* (connecting cortex → striatum → external → GPi and → STN / substantia nigra with many interconnections among regions), which we termed the *main axis of the basal ganglia* in the introduction. On the second hand, the pallidum is traversed *orthogonally* to the main axis by the pallidothalamic projections (which are thought to form a continuum and include fasciculus and ansa lenticularis ^15,51^. This leads us to the second stage of our model, which aimed at modeling exactly these differential fiber systems.

To briefly summarize tract-based findings again: Modulating posterior pallidosubthalamic/nigral fibers (main axis) accounted for optimal outcomes in cervical dystonia, while modulating pallidothalamic fibers for generalized dystonia – both with the common projection back to the thalamus. While the former would be thought to map more strongly onto the indirect pathway (with the STN) and/or use the nigra as output structure, the latter would primarily implement the GPi as output structure. We especially deem the latter finding crucial since, as mentioned, the pallidothalamic bundles traverse the whole pallidum and could integrate information from all pallidal regions. Hence, speculatively, disruption at this system could lead to more generalized symptoms not limited to specific body parts. In turn, widely parallel fibers radially projecting from the pallidum through the internal capsule (i.e., extending the parallel striatopallidofugal fibers), could be affected in a more symptom and somatotopy-specific fashion. We must emphasize that this notion is and remains speculative and could only be confirmed and deliberately tested in animal models and axon-specific modulation strategies (e.g., using optogenetics). A more intuitive second finding was those fibers of passage projecting from head/cervical vs. generalized somatotopic regions of somatomotor cortex differentially accounted for variance in outcomes in the respective dystonia type. Alterations in somatomotor cortex, specifically in plasticity of the sensory cortices, have been proposed to play a crucial pathophysiological role in dystonia ^52–57^. Recently, Corp et al. applied lesion network mapping to investigate shared networks of stroke lesions that led to cervical dystonia, which again attributed a specific role to somatosensory cortex ^58^. Hence, our findings that successful treatment of cervical dystonia – at least by somatotopic domain – maps to connections to specifically the head/cervical zones and generalized dystonia to the full somatotopic domain of somatomotor cortex could form crucial additional support for pathophysiological involvement of the somatomotor cortex. In this context, however, it is crucial to emphasize that direct projections from cortex to pallidum are not classically described in the gross-anatomical literature and hence, may at least not exist in large numbers. While reports about such direct connections (also between an aforementioned peripallidal site around the GPi and cortex ^48^) have been described using robust methods that are not prone to false-positive connections ^59^ (for an overview also see fig. 25 in ^60^), the largest proportion of cortical input to the pallidum is transmitted via the massive projection of the striatopallidofugal bundle ^46^. Hence, the direct cortical connections our analysis revealed *could* be truly pathophysiologically relevant. Alternatively, they could express the specific region of the pallidum that would likely receive cortico*striatal* input from the same cortical regions, given the orderly fashioned organization of the whole cortico-basal ganglia thalamic loops ^29,61,62^.

On a whole-brain level that could involve polysynaptic connections, a region of longstanding interest for the pathophysiology of dystonia is the cerebellum ^63^. This leads us to the final stage of our model which involved the broad-lense view of whole-brain functional networks.

### Network-level stimulation model

On a whole brain level, the aforementioned study by Corp et al. revealed that lesions which led to cervical dystonia would be positively connected to the cerebellum and negatively to the sensory cortex ^58^. Crucially, among the few regions with connections that did account for symptom improvement following DBS regardless of dystonia type were exactly these two regions with the same signs as described by Corp et al. (*Agreement map* in figure 7). Specifically, the positive connections to wider parts of the cerebellum were positively associated with optimal outcomes in both cervical and generalized dystonia. Functional involvement of the cerebellum could be mechanistically implemented by malfunctions of the sodium-potassium pump in cerebellar Purkinje neurons ^64^ with ouabain blocks of the pump leading to dystonic symptoms in mouse models ^65^. Clinical reports involve disappearance of dystonic symptoms after cerebellectomy ^66^ and substantial antidystonic effects after DBS to the cerebellum – even after failed bilateral pallidotomy and intrathecal baclofen therapy ^67^. Hence, while dystonia has traditionally been regarded as a basal ganglia disorder, enough certainty has accumulated that its pathophysiology involves cortico-ponto-cerebello-thalamo-cortical loops, as well. While basal ganglia input to the thalamus maps to matrix cells, which diffusely project to apical cortical layers, cerebellar thalamic input predominantly maps to core cells that focally project to basal dendrites of layer five cortical neurons ^62,68^. This has led to the notion that the cerebellar function in movement embodies a system to automatize certain types of movements after motor learning ^62,69^. In the thalamus, cerebellar and basal ganglia input is integrated to orchestrate cortical activity *and plasticity* (^62^; due to transition to burst-mode firing of cortical neurons in case of simultaneous activation of apical and basal dendrites, ^70^). Our results show supporting evidence for the involvement of exactly these pallidothalamic projections for the case of generalized dystonia (and multisynaptic involvement of the cerebellum). In case of cervical dystonia, results suggest an indirect connection via subthalamic nucleus but similar polysynaptic involvement of the cerebellum. Alternatively, the pallidosubthalamic fibers could represent pallidonigral projections, which are impossible to differentiate by means of neuroimaging given their intertwined course within Edinger’s comb system. In the latter case, nigrothalamic projections and multisynaptic cerebellar involvement would constitute an analogous finding to the one described in generalized dystonia.

### Limitations

Multiple limitations on various levels apply to the present study. First and foremost, the retrospective nature of the study should be emphasized, as well as the analysis of the same multi-center cohort in the study by Reich et al. ^6^. Still, the sample constitutes the largest cohort studied by means of DBS imaging, to date, and we believe that studying it with different methods and approaches will be beneficial for treating dystonia going forward. Along the same lines, the sample involves heterogeneous imaging datasets as well as clinical records that were acquired during clinical routine. This adds to the inherent imprecision of DBS electrode localizations which can be substantial and largely depend on imaging quality ^37^, however could also be seen as a strength since variability in the data may lead to more robust findings not overfit to data from a specific surgeon/center. We applied a modern imaging pipeline that has been specifically developed to localize DBS electrodes, including multispectral normalizations ^29^, manual warpfield refinements ^27^ and brain shift correction ^20^, as well as phantom-validated electrode localizations ^28^. Automatic segmentations of the GPi derived with the pipeline rivaled the precision of manual expert segmentations in a study that investigated multiple nonlinear registration approaches for the subcortex ^25^ and a second study that confirmed results ^26^. Furthermore, manual refinement of registrations was applied in a labor-intensive patient-by-patient process to ensure precise fit between the atlas model of the pallidum and the patient-specific MRI data ^27^. Still, a certain degree of imprecision is inherent to this process and must be acknowledged.

Third, on the tract-level, the accuracy and anatomical validity of the basal ganglia pathway model is an important condition to interpret our results. Here, slight misrepresentations of the anatomical detail of implemented tracts could have large impact on results. For instance, the *intrapallidal* course of the pallidothalamic output fibers would likely play a role in segregating results for cervical and generalized dystomia. While the pathway model has been curated by world-reknowned basal ganglia anatomists ^17^ and constitutes the best anatomical model our field currently has, it has been indirectly defined in humans (and, for instance, was largely informed by macaque tracer studies ^71^) and – as any normative atlas resource – does not account for individual variability. Hence, interpretation of especially the tract-level results of the present study should be seen as a function of anatomical validity of the tract atlas.

Fourth, the biophysical model electrical effects on the tissue was modeled in a comparably simplified manner, where more advanced concepts have been introduced, in the past ^35,36,72^. While more advanced biophysical modelling options have now been introduced as open-source and interfaced within the Lead-DBS software applied here, ^73^, as mentioned in our methods section, in the present study, the choice of a simple model was indeed deliberate. Modeling an electric field is a simpler engineering task which is often followed by modeling *biology*, i.e., the response of neurons and axons. A downside of the latter approach is the necessity to impose a large set of assumptions. Instead, here, we did apply concepts that would not be susceptible to the exact relationship between stimulation amplitude and the degree of neuromodulation on the axonal populations surrounding the electrodes. Instead, the model (concretely implemented by means of mass-univariate rank-correlations) would yield maximal weights for any type of monotonically increasing relationship between the two.

### Conclusions

We report evidence that cervical vs. generalized dystonia responds optimally to neuromodulation of a specific set of pallidofugal and pallidothalamic connections and that treatment effects involve indirect connections with the cerebellum and somatomotor cortex. Specific optimal stimulation sites in the pallidum map to somatotopic representations of the nucleus, with the optimal stimulation site for cervical dystonia mapping to its cervical functional zone. We construct a model that involves local, tract-and network-based components that explain significant amounts of clinical variance following DBS to the pallidum.

## Data Availability

The DBS MRI datasets generated during and analyzed during the current study are not publicly available due to data privacy regulations of patient data but are available from the corresponding author on reasonable request. All code used to analyze the dataset is available within Lead-DBS /-Connectome software (https://github.com/leaddbs/leaddbs). 

## Acknowledgements

A.H. was supported by the German Research Foundation (Deutsche Forschungsgemeinschaft, Emmy Noether Stipend 410169619 and 424778381 – TRR 295) as well as Deutsches Zentrum für Luft-und Raumfahrt (DynaSti grant within the EU Joint Programme Neurodegenerative Disease Research, JPND). A.H. is participant in the BIH-Charité Clinician Scientist Program funded by the Charité – Universitätsmedizin Berlin and the Berlin Institute of Health.

## Author contributions

A.H. conceptualized the study, developed the software pipeline used, analyzed data, and wrote the manuscript. M.R. acquired data and critically revised the manuscript. S.E. conceptualized the study, analyzed data, and critically revised the manuscript. B.A-F. analyzed data and critically revised the manuscript. N.L. developed the software pipeline used and critically revised the manuscript. I.H. analyzed data, created figures and critically revised the manuscript. F.L., J.R., S.P., J.R., F.W., N.G.P., K.W., R.C.N., L.S., V.M., M.W., G-H. S., V.C., P.M., W.P., W.E., A-K. H., C.M., V.S., I.U.I., J.K.K., G.D. and J.V. acquired data and critically revised the manuscript. A.A.K. conceptualized the study and wrote the manuscript.

## Materials & Correspondence

Correspondence and material requests should be addressed to Andreas Horn.

## Supplementary material

### Further anatomical considerations

The following considerations went into forming the hypothesis of the present manuscript, that have only briefly been touched upon in the introduction and **figure 1** (main text): Together with the reticular part of the substantia nigra (SNr), the GPi constitutes the *output ganglion* of the basal ganglia, feeding *cortical* signals that arrived at the striatum and external pallidum (GPe) via the thalamus back to the cortex ^74–76^. This loop model has become fundamental in our understanding of movement disorders such as dystonia, Parkinson’s Disease and other *‘basal ganglia disorders’*. Multiple refinements of this model have been proposed since ^16,60,77^ and converging models have been developed in parallel within the basal ganglia and reinforcement learning fields (see **Figure 1** A for an example). These models have often been defined in form of boxes and arrows to show *functional* interactions between structures. Concepts such as the direct, indirect and hyperdirect pathways ^78,79^ have become fundamental for our understanding ^77^, but are primarily functional concepts. While their anatomical correlates have been investigated ^14,78,80–82^, in the larger body of functional basal ganglia studies, less focus has been put on the exact tracts that implement direct and indirect pathways (including their projections back to thalamus and cortex).

To derive at a circuit-based hypothesis of DBS in anatomical space, a translation to specific anatomical structures is necessary (**Figure 1** B). One component that has been well characterized on an anatomical level is the hyperdirect pathway. Parts of descending glutamatergic projections from cortex to the motor pattern initiator / generator networks in the brainstem ^60^ send axon-collaterals to multiple subcortical regions such as the subthalamic nucleus (STN) which functionally define what we call the hyperdirect pathway ^78,79,82^. Anatomical correlates of the direct and indirect pathways are intermixed and implemented via i) the striatopallidofugal bundle and ii) Edinger’s comb system. The former, a massive white-matter structure that traverses the striatum (in form of Wilson’s pencils ^83,84^) and both parts of the pallidum radially while partly rewiring in the laminae externae, internae and accessoriae (**Figure S1 B**), hence partly forming indirect pathway (synapsing within GPe) and direct pathway (projecting from striatum directly to GPi). Another crucial functional component of the indirect pathway is the STN which is connected via an extension of the striatopallidofugal system that traverses the internal capsule orthogonally and forms part of the comb system of Edinger ^84,85^. Macroscopically, the same structure also shows connections between striatopallidal regions and the SNr. Finally, pallidothalamic fibers are traversing the pallidum in *orthogonal* fashion to the striatopallidofugal system. One reason could be that terminal fields of pallidal neurons that integrate information from striatal domains are arranged in a disc-like fashion orthogonal to the striatopallidofugal bundle (**Figure S1**). Crucially, these disc-like receptive fields of pallidal neurons are of *fixed size* and hence integrate information from increasingly larger striatal (and hence cortical) zones medially ^14,60,80^. Hence, increasingly medial pallidal neurons (even within the GPi) seem to form the largest integrator hub regions within the pallidum. Finally, a quite exclusive property of the STN is that it has no known direct efferents to the thalamus, i.e., its feedback to the loop is indeed quite indirect (while the GPe does project to the thalamus, directly; ^60^).

**Figure S1:**
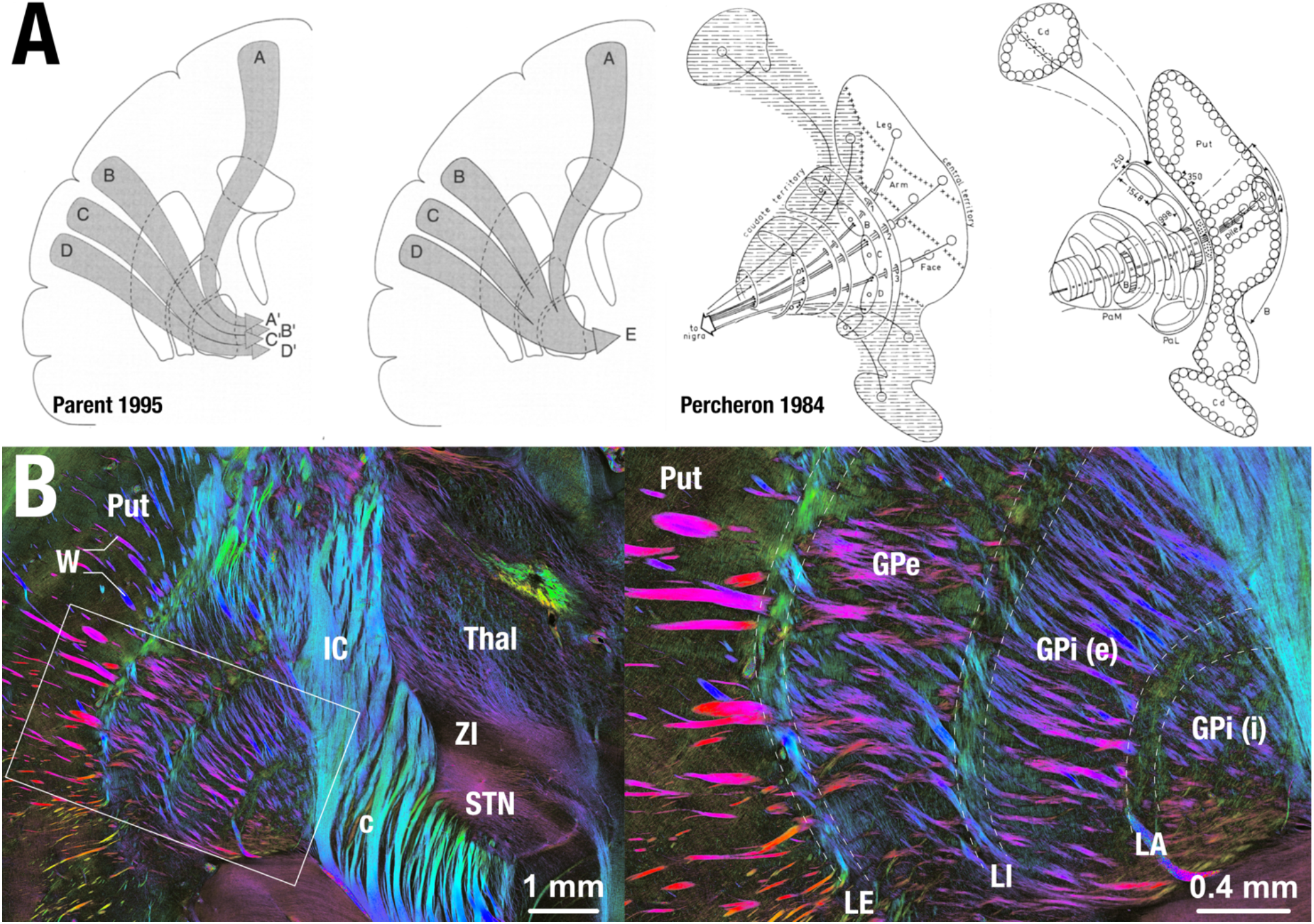
Anatomical considerations. A) As two basic frameworks, the parallel loops and funnel concepts were reviewed by Parent et al. 1995. The concepts are not competing, rather, both are partly implemented by the corticostriatal and striatopallidofugal system of the basal ganglia. Information from different cortical sites (A-D) are partly kept segregated (leading to information sites in A’-D’) but also integrated (leading to a joint information site E). Hence, the basal ganglia both integrate information from cortical sites but partly keep information separate. This was nicely illustrated by Percheron et al. 1984, showing receptive fields of pallidal neurons that are organized in disk-like fashion orthogonal to incoming striatal projections. Of note, size of each disk is constant, leading to higher degrees of integration in increasingly medial parts of the pallidum. Panels of original publications reproduced with permission. B) Polarized Light Imaging data acquired in the vervet monkey shows multiple stages of data compression and rewiring but also shows parallel organization of incoming loops. Image courtesy by Markus Axer and Karl Zilles, data from ^86^.

As mentioned, pallidothalamic projections – anatomically realized by the ansa and fasciculus lenticularis which merge within Forel’s field H1 to form the fasciculus thalamicus ^51^ – traverse the GPi *parallel* to its maximal extent and predominantly project to the pallidal part of the ventroanterior nucleus of the thalamus (VAp; (Ilinsky *et al*., 2018)). An older model claimed that the fasciculus lenticularis would integrate projections from the external part of the GPi while the ansa the ones from the internal part of the GPi. However, this model has been revised and it was suggested that the two tracts rather form a joint functional unit ^15,51^. We will adopt this view here, i.e., subsummize ansa and fasciculus lenticularis as *pallidothalamic projections*.

DBS in our cohort was applied to a single node of this complex network: the GPi. With a GPi-centric view in mind, the network can be dramatically simplified to two fiber systems that traverse in *quasi-orthogonal direction* to each other – which is exactly what we aimed to leverage in the present study.

